# Parametric Survival Analysis of Long COVID Among Hospitalized Patients in Zambia: A Retrospective Cohort Study on the Time to Symptoms Resolving

**DOI:** 10.1101/2025.05.05.25327037

**Authors:** Warren Malambo, Mutale Sampa-Kawana, Duncan Chanda, Sombo Folowshi, Jonas Z. Hines, Patrick Kaonga

## Abstract

**Background:** Long COVID refers to the continuation or development of new symptoms within three months after acute SARS-CoV-2 infection, with these symptoms lasting for at least 2 months. Despite studies on COVID-19 sequelae, gaps remain in understanding the temporal dynamics of COVID-19 symptom resolving. This knowledge is crucial for treatment planning and setting realistic recovery expectations. Using a cohort of patients hospitalized in Zambia for COVID-19, the objective of this study was to evaluate resolution of COVID-19-related symptoms over time and associated factors.

**Methods:** We conducted a retrospective cohort study of persons discharged after COVID-19 hospitalization and presenting for follow-up care in 13 specialized clinics in Zambia from August-2020 to December-2022. All participants were hospitalized with acute COVID-19. Severe acute COVID-19 was defined as hospitalization further requiring supplemental oxygen therapy, ICU stay, and/or treatment with steroids/remdesivir. We evaluated time-to-symptoms resolving (i.e., survival time) as our primary outcome. We estimated symptoms resolution incidence rate, median survival time (onset-to-resolution), associated factors, and changes in the hazard of symptoms resolving using survival analysis.

**Results:** Of the 823 study participants, 616 (84.3%) had severe acute COVID-19 illness, half (50.6%) were female, and median age was 54 (interquartile range [IQR]: 43-64) years. Nearly three-quarters (597, 72.5%) had their symptoms resolved at a median survival time of 51 (IQR: 34-104) days. A majority of participants (59.4%) had baseline comorbidities, 16.6% had prior vaccination with ≥1 COVID-19 vaccine dose, and median acute hospitalization duration was 8 (IQR: 4-16) days. Persistent COVID-19 symptoms resolved at a rate of 12.2 per 1,000 person-days. Severe acute COVID-19 (adjusted hazard ratio [aHR]: 0.65, 95% CI: 0.41-0.88) and having ≥1 comorbidity (adjust time ratio [aTR]: 1.86, 95% CI: 1.60-2.17) was associated with slower resolution. Infection during the Omicron-predominant period compared to Alpha (aHR: 4.05, 95% CI: 1.27-12.9) and prior vaccination before illness (aTR: 0.50; 95% CI: 0.26-0.94) were associated with faster symptom resolution. The changes in the hazard rate of symptoms resolving was non-linear, it instead increased to a peak rate of 2.14% per day at 20 days and then decreased.

**Conclusion:** An increasing then decreasing hazard rate trajectory meant that COVID-19 symptoms resolved relatively faster in the first month. The median survival time occurring nearly a month after peak hazard rate could suggest some patients who do not initially improve in the first month of their post-acute infection were less likely to resolve afterwards. This may likely help in treatment planning and in providing persons with persistent COVID-19 symptoms realistic expectations about their recovery process.

## INTRODUCTION

Long COVID, a post-viral syndrome, may occur after SARS-CoV-2 infection and affect multiple-organ systems. While some individuals may fully recover shortly after a SARS-CoV-2 infection, others experience persistent symptoms that require ongoing management and support. Long COVID presents in a broad spectrum of symptoms but the most common may include fatigue, shortness of breath, headaches, myalgias, cough and cognitive changes [1]. WHO defines long COVID as the continuation or development of new symptoms 3 months after the initial SARS-CoV-2 infection, with symptoms lasting for at least 2 months and with no other explanation [2]. Despite this definition, a lack of consensus on the clinical presentation of long COVID or a diagnostic test or biomarker for long COVID complicates the categorization and analysis of its sequelae.

A framework, however, suggests that the spectrum of long COVID conditions may be delineated as: i) a group of persisting symptoms that interfere with the ability to function at home or at work and are not explained by major organ injury; ii) new onset of major diseases including diabetes mellitus, cardiovascular diseases, stroke, and pulmonary failure; and iii) tissue injury of multiple organs that has the potential to lead to long-term organ dysfunction [3]. This suggests long COVID may encompass a spectrum of conditions ranging from mild but disabling symptoms (without organ damage) to more severe cases involving the development of serious diseases or actual physical damage to organs.

Evidence on long COVID in Africa has steadily increased despite limited access to health-care services by patients, and inadequate diagnostics and clinical data [4]. The prevalence is estimated to range between 2% to 86% [4-8]. It varies between 10-30% in patients with mild COVID-19, 50-70% in acute COVID-19 hospitalized patients and 10-12% among vaccinated cases [9-13]. Associated factors include older age (≥40 years), underlying comorbidity, severe COVID-19 disease, and admission to intensive care [4, 14-16]. Studies have further shown that vaccination is associated with a reduced likelihood of severe acute COVID-19 and long COVID [16-18]. Long COVID impacts on patients’ quality of life, employment, and income due to diminished functional status or protracted path to recovery [19, 20].

Although a number of studies in Zambia have reported COVID-19 sequelae, the knowledge gap still exists around the temporal dynamics of COVID-19 symptoms resolving [21-28]. Such information is vital in treatment planning for long COVID and in providing patients with realistic expectations about their post-acute COVID-19 recovery process. In the acute phase of COVID-19, patients hospitalized for COVID-19 in Zambia had a median recovery time of 12 days [29]. In the post-acute phase, it’s reported that some individuals still had symptoms at nearly two months of follow up [15, 27]. None of these studies, however, report on the trajectory or temporal dynamics of long COVID-19 resolution. In this study, we analyzed persistent COVID-19 symptoms to determine the incidence rate, median survival time, factors associated with time-to-resolving, and changes in hazard rate of symptoms resolving.

## METHODS

### Study setting

In August 2020, the Zambian Ministry of Health launched 13 referral clinics to provide post-acute COVID-19 (PAC-19) care for individuals recovering from COVID-19. PAC-19 clinics were established to provide comprehensive care given the unknown long-term sequalae of COVID-19 at the time [30]. Individuals were also evaluated for persistent or new symptoms to assess whether these were stable, worsening, and/or new since this could reflect development of late complications of COVID-19. The evaluation was through clinicians’ review of systems using standardized forms.

### Study participants

At a first clinic visit, patients’ demographic and medical history were recorded on standardized paper forms which were routinely abstracted into a REDCap electronic database. Clinicians assessed for current post-COVID symptoms and comorbidities (e.g., hypertension, diabetes, cardiovascular disease, HIV or TB) coupled with physical examination and laboratory investigations including COVID-19 test, if required. A review of systems included assessment for general, cardiovascular, pulmonary, gastrointestinal, urinary, neurologic, musculoskeletal, ENT (ear, nose and throat), mental health, and dermatologic symptoms.

Patients attended up to five follow-up visits based on an individuals’ parameters and clinicians’ recommendation. Follow-up schedules were individualized for physical exams and, when necessary, reviews were also made via phone calls for some individuals. Discharge criteria from PAC-19 clinics was based on symptoms resolution (i.e., no reported COVID-19 symptoms by patients) and normal physical/laboratory parameters as assessed by clinicians. Patients with new medical conditions onset (e.g., hypertension, diabetes mellitus, cardiovascular diseases, stroke, or pulmonary failure) or requiring further specialist care (e.g., physiotherapy, cardiology, endocrinology, nephrology, mental health, and pulmonology) were referred for further follow-up at appropriate units.

### Study design

We implemented a retrospective cohort study design. Participants were included in the study if they presented in the PAC-19 clinics between 20-Aug-20 (study start date) and 31-Dec-22 (study end date), and had their information captured in a REDCap (Research Electronic Data Capture) electronic database [31]. Data from REDCap was initially accessed on 5-Jan-2024 and analyzed for this study. To ensure the cohort was similar on all baseline characteristics, outpatients during acute COVID-19 who presented at PAC-19 clinics were excluded from the study due to differences in clinical course management.

### Study variables

The primary exposure variable was hospitalization within COVID-19 during the study period. This group was further defined as those with “mild” or with “severe” acute COVID-19. Severe acute COVID-19 disease was defined as an episode that required supplemental oxygen therapy or intensive care unit stay or treatment with steroids/or remdesivir. The mild COVID-19 cohort were admitted during acute COVID-19 infection but did not require supplemental oxygen therapy or intensive care unit stay or treatment with steroids/or remdesivir. The outcome of interest was whether the COVID-19 symptoms resolved (coded as 1 if participants experienced symptoms resolution at any time point during the study or 0 if they were right censored) and the time it took to symptom resolving or censor. Participants were categorized as censored if their symptoms didn’t resolve by their last follow-up time point or were lost to follow-up (LTFU) but has at least 1 PAC-19 clinic visit (Supp. Fig 1). Time-to-resolution was the time interval, in days, from the date of COVID-19 diagnosis to the date of symptom resolving or censor.

Other study covariates were sex, age group, dominant SARS-CoV-2 variant in Zambia at time of diagnosis, presence of baseline comorbidities, new onsite comorbidities, length of hospitalization, and vaccination status. The classification of SARS-CoV-2 dominant variant was based on data from Zambia’s genomic surveillance system submitted to the Global Initiative on Sharing All Influenza Data (GISAID) [32]. Per GISAID, the Alpha variant was dominant from 3-March-2020 to 21-September-2020, the Beta variant up to 19-March-2021, the Delta variant up to 12-December-2021 and the Omicron variant was still dominant at the study end date of 31-December-2022. Vaccination status was classified as unvaccinated, prior vaccination (defined as 1 or more dose of an approved COVID-19 vaccine) for those vaccinated before their acute COVID-19, and vaccinated during PAC-19 clinic follow up. This was based on vaccination records, when available, or patients’ self-reported status.

### Bias

To control for selection bias, participants presenting for follow-up that met the inclusion criteria were in the study to ensure representativeness of the target population. Lost to follow-up bias was addressed by including time points preceding censoring. Measurement bias due to misclassification of long COVID was addressed by excluding patients from outpatient episodic who may have presented for care during acute COVID-19 infection. Confounding bias was controlled for by including other covariates in addition to the exposure variable at multivariable analysis.

### Sample Size

The required minimum sample size was 396 based on the cohort studies sample size formula, a previously reported long COVID prevalence assumed at 41% among a severe acute COVID-19 cohort and at 17% among mild COVID-19 cohort [33-35]. We further assumed a ratio of severe COVID-19 to mild COVID-19 of two. We nonetheless considered a full enumeration of 823 participants that met the study inclusion criteria for representativeness.

### Statistical analysis

Using descriptive statistics, we assessed baseline demographic and clinical characteristics of study participants. Frequencies with proportions were presented for categorical covariates and medians with the interquartile range (IQR) for non-normally distributed numeric covariates. We reported measures of association to the outcome using the Pearson chi-square, Kruskal-Wallis rank sum, and Fishers’ exact tests p-value. Adjustment for multiple pairwise comparisons between the five clinical visits were done using Bonferroni correction.

At inferential statistics, we fitted the Kaplan-Meier estimator to compute the total person time at risk, symptom resolution incidence rate, and the median survival time. Survival was defined as the probability of not having experienced symptoms resolution up to a certain time. Time was the number of days to symptoms resolution or censor. We assessed factors associated with time-to-resolution of persistent COVID-19 symptoms using a Cox proportional hazard model. In this model, we checked for the proportional hazard assumption by plotting the Schoenfeld residual against the transformed time to check linearity. We fitted a stratified Cox proportional hazard model to control for violation of proportionality assumption by the covariates presence of comorbidity and hospital length of stay. The hazard was defined as the instantaneous risk of experiencing long COVID resolution at a specific moment in time, given that the symptoms had survived up to that time.

Since the data had a longitudinal structure, i.e., repeated observation of symptoms over clinical review visits, we implemented a parsimonious approach to fitting a stratified Cox proportional hazard model to account for the supposed dependence structure. First, we fitted a Cox proportional hazard model without taking account of the supposed dependence or longitudinal structure, followed by a model with sandwich (robust) estimator, and finally a mixed effects Cox proportional hazard model. The random term in the mixed effects model was the patient since multiple observations were longitudinally made for each patient. The mixed effects Cox proportional hazard model was preferred based on the log likelihood of observing the survival data and the lowest Akaike Information Criteria (AIC).

To explore the temporal dynamics of persistent COVID-19 symptoms resolving (i.e., hazard rate which is the instantaneous rate of occurrence of symptom resolving in a given day), we further fitted a parametric survival model which assume that survival time follow a specific trajectory. To select the underlying parametric form or, simply, the underlying hazard rate distribution that symptoms revolving survival time may have followed, we compared 5 distributions (exponential, Weibull, log-logistic, log-normal, and generalized gamma). The generalized gamma distribution (Supp Fig. 2) was the preferred parametric form based on the log-likelihood and AIC values. We reported the accelerated failure time (AFT) under the assumed parametric distribution, i.e., whether the presence of a covariate led to shorter or longer symptoms survival times (when linearly related on the logarithm scale). All analysis were done in R version 4.4.3 and statistical significance was considered at p<0.05.

### Ethics

Ethical clearance (waiver for informed consent) was obtained from the University of Zambia Biomedical Research Ethics Committee (Ref No. 3482-2022), and approval to conduct the study was obtained from the Zambia National Health Research Authority (Ref No: NHRA0000011/09/02/2023). The investigators did not interact with study participants but only had access to anonymized data.

## RESULTS

Of the 823 study participants, slightly over half were female (50.6%) and the median age was 54 (interquartile rang [IQR]: 43-64) years (Table 1). Close to one-third (n=303, 36.8%) had their symptoms resolved by their first visit to a PAC-19 clinic with reference to the date of COVID-19 diagnosis. Overall, 597 (72.5%) participants had their COVID-19 symptoms resolve during the five follow up visits. Of the total number of participants, 70.1% (501) were diagnosed with COVID-19 when Delta was the dominant variant. Four hundred thirty-seven (57.4%) participants had baseline comorbidities, with the most frequently reported underlying medical conditions being hypertension (40.6%), diabetes mellitus (16.2%), HIV (15.6%) and cardiovascular disease (6.6%). Of all patients, 115 (14%) had new medical conditions onset at the time of acute COVID-19 that included diabetes (n=59, 51.3%), hypertension (n=50, 43.5%) and other medical conditions (n=19, 16.5%).

**Table 1:** Baseline Demographic and clinical characteristics of study participants who presented for care in specialized PAC-19 clinics in Zambia, Aug. 2020 to Dec. 2022 (N=823)

| Characteristic | Longitudinal Follow up Review visits Attendance |  |  |  |  | p-value<sup>1</sup> |
| --- | --- | --- | --- | --- | --- | --- |
|  | Visit 1 (823)<br>n (%) | Visit 2 (374)<br>n (%) | Visit 3 (170)<br>n (%) | Visit 4 (83)<br>n (%) | Visit 5 (36)<br>n (%) |  |
| **COVID-19 Symptoms Survival status** |  |  |  |  |  | **<0.001<sup>P</sup>** |
| Censored | 520 (63.2) | 218 (58.3) | 99 (58.2) | 40 (48.2) | 12 (33.3) |  |
| Resolved | 303 (36.8) | 156 (41.7) | 71 (41.8) | 43 (51.8) | 24 (66.7) |  |
| **Sex** |  |  |  |  |  | **0.955<sup>P</sup>** |
| Male | 400 (49.4) | 175 (47.4) | 81 (48.5) | 41 (49.4) | 16 (44.4) |  |
| Female | 410 (50.6) | 194 (52.6) | 86 (51.5) | 42 (50.6) | 20 (55.6) |  |
| (Missing) | 13 | 5 | 3 | 0 | 0 |  |
| **Age (years)** |  |  |  |  |  | **<0.001<sup>W</sup>** |
| Median (IQR) | 54 (43, 64) | 57 (47, 66) | 59 (49, 68) | 61 (50, 69) | 61 (52, 69) |  |
| (Missing) | 11 | 2 | 1 | 1 | 1 |  |
| **Age group (years)** |  |  |  |  |  | **<0.001<sup>F</sup>** |
| <29 | 55 (6.8) | 9 (2.4) | 3 (1.8) | 1 (1.2) | 1 (2.9) |  |
|  | Visit 1 (823)<br>n (%) | Visit 2 (374)<br>n (%) | Visit 3 (170)<br>n (%) | Visit 4 (83)<br>n (%) | Visit 5 (36)<br>n (%) |  |
| 30-39 | 89 (11.0) | 31 (8.3) | 13 (7.7) | 4 (4.9) | 1 (2.9) |  |
| 40-49 | 175 (21.6) | 82 (22.0) | 29 (17.2) | 15 (18.3) | 6 (17.1) |  |
| 50-59 | 203 (25.0) | 97 (26.1) | 43 (25.4) | 16 (19.5) | 5 (14.3) |  |
| 60+ | 290 (35.7) | 153 (41.1) | 81 (47.9) | 46 (56.1) | 22 (62.9) |  |
| (Missing) | 11 | 2 | 1 | 1 | 1 |  |
| **Dominant SARS-CoV-2 variant at diagnosis<sup>2</sup>** |  |  |  |  |  | **<0.001<sup>F</sup>** |
| Alpha | 39 (5.5) | 21 (6.2) | 17 (11.0) | 7 (8.8) | 3 (8.6) |  |
| Beta | 82 (11.5) | 51 (15.1) | 26 (16.8) | 9 (11.3) | 2 (5.7) |  |
| Delta | 501 (70.1) | 249 (73.9) | 109 (70.3) | 61 (76.3) | 30 (85.7) |  |
| Omicron | 93 (13.0) | 16 (4.7) | 3 (1.9) | 3 (3.8) | 0 (0) |  |
| (Missing) | 108 | 37 | 15 | 3 | 1 |  |
| **Presence of comorbidities<sup>3</sup>** |  |  |  |  |  | **<0.001<sup>P</sup>** |
| No | 346 (42.6) | 136 (36.6) | 52 (30.8) | 22 (26.5) | 7 (19.4) |  |
| Yes | 467 (57.4) | 236 (63.4) | 117 (69.2) | 61 (73.5) | 29 (80.6) |  |
| (Missing) | 10 | 2 | 1 | 0 | 0 |  |
| **New medical conditions onset<sup>4</sup>** |  |  |  |  |  | **0.918<sup>P</sup>** |
| No | 708 (86.0) | 316 (84.5) | 144 (84.7) | 69 (83.1) | 31 (86.5) |  |
| Yes | 115 (14.0) | 58 (15.5) | 26 (15.3) | 14 (16.9) | 5 (13.5) |  |
| **Hospitalization duration (days)** |  |  |  |  |  | **0.003<sup>K</sup>** |
| Median (IQR) | 8 (4, 16) | 9 (6, 19) | 11 (6, 20) | 10 (6, 19) | 9 (6, 19) |  |
| (Missing) | 228 | 104 | 43 | 15 | 5 |  |
| **Hospital length of stay (days)** |  |  |  |  |  | **0.008<sup>F</sup>** |
| 1-3 | 110 (18.5) | 26 (9.6) | 10 (7.9) | 4 (5.9) | 2 (6.5) |  |
| 4-7 | 162 (27.2) | 73 (27.0) | 33 (26.0) | 19 (27.9) | 10 (32.3) |  |
| 8-14 | 157 (26.4) | 81 (30.0) | 39 (30.7) | 21 (30.9) | 9 (29.0) |  |
| ≥15 | 166 (27.9) | 90 (33.3) | 45 (35.4) | 24 (35.3) | 10 (32.3) |  |
| (Missing) | 228 | 104 | 43 | 15 | 5 |  |
| **Severe acute COVID-19<sup>5</sup>** |  |  |  |  |  | **0.216<sup>F</sup>** |
| No | 115 (15.7) | 39 (11.3) | 17 (10.7) | 9 (11.1) | 4 (11.4) |  |
| Yes | 616 (84.3) | 307 (88.7) | 142 (89.3) | 72 (88.9) | 31 (88.6) |  |
| (Missing) | 92 | 28 | 11 | 2 | 1 |  |
| **Vaccination status<sup>6</sup>** |  |  |  |  |  | **<0.001<sup>P</sup>** |
| Unvaccinated | 625 (83.4) | 273 (75.6) | 120 (71.9) | 44 (53.0) | 14 (38.9) |  |
| Prior vaccination | 124 (16.6) | 49 (13.6) | 13 (7.8) | 9 (10.8) | 3 (8.3) |  |
| Vaccination during follow up | 0 | 40 (11.1) | 35 (21.0) | 30 (36.1) | 19 (52.8) |  |
| (Missing) | 74 | 13 | 3 | 0 | 0 |  |
| **Time since COVID-19 diagnosis (days)** |  |  |  |  |  | **<0.001<sup>K</sup>** |
| Median (IQR) | 30 (20, 42) | 51 (37, 70) | 75 (57, 99) | 92 (74, 115) | 110 (97, 143) |  |
| (Missing) | 109 | 37 | 15 | 3 | 1 |  |

1. Bolded p-values are significant at p<0.05 and were Bonferroni adjusted for multiple comparisons. Tests performed included Pearson Chi-square test (P), Kruskal-Wallis rank sum test (K), and Fishers' Exact Test (F).
2. Dominant SARS CoV-2 variants classification based on GISAID (2023) rather than sequenced specimens from study patients
3. Pre-existing comorbidity: hypertension, diabetes, cardiovascular disease, cancer, immunosuppression, chronic lung, kidney, and liver diseases, obesity, HIV and TB.
4. New medical conditions onset included diabetes, hypertension, deep vein thrombosis/pulmonary embolism and kidney injury/disease
5. Composite variable for acute COVID-19 episodic that required supplemental oxygen therapy, intensive care unit stay or treatment with steroids/remdesivir
6. Vaccination status was a time vary covariate. Some participants initially unvaccinated got vaccinated during follow up.

During acute COVID-19 hospitalization, 616 (84.3%) participants had severe disease, and the median hospital length of stay was 8 (IQR: 4-16) days. Of all participants, 124 (16.6%) were vaccinated (at least ≥1 dose) prior to COVID-19 infection and during follow up, the number of vaccinated participants more than doubled to 320 (38.9%). Of those vaccinated, 140 (43.8%) received AstraZeneca (AZD1222), 152 (47.5%) Johnson and Johnson’s Janssen (Ad26.COV2.S), 27 (8.4%) did not know the vaccine type they received and 1 (0.3%) received Pfizer-BioNTech (BNT162b2). The median time since COVID-19 diagnosis patients were followed up was 42 (IQR: 26-72) days while the maximum follow-up time was 260 days.

Overall, the most prevalent persistent COVID-19 symptom classes participants presented with included pulmonary (cough 17.2%, chest pain 10.3% and shortness of breath 7.8%), general symptoms (fatigue 15.6%), neurologic (headache 7.3%), cardiovascular (palpitations 6.0%) and musculoskeletal (myalgia 5.0% and joint aches/pain 4.6%) related. The proportions of reported symptoms displayed a declining trend across clinical visits (Fig 1).

**Fig. 1:**
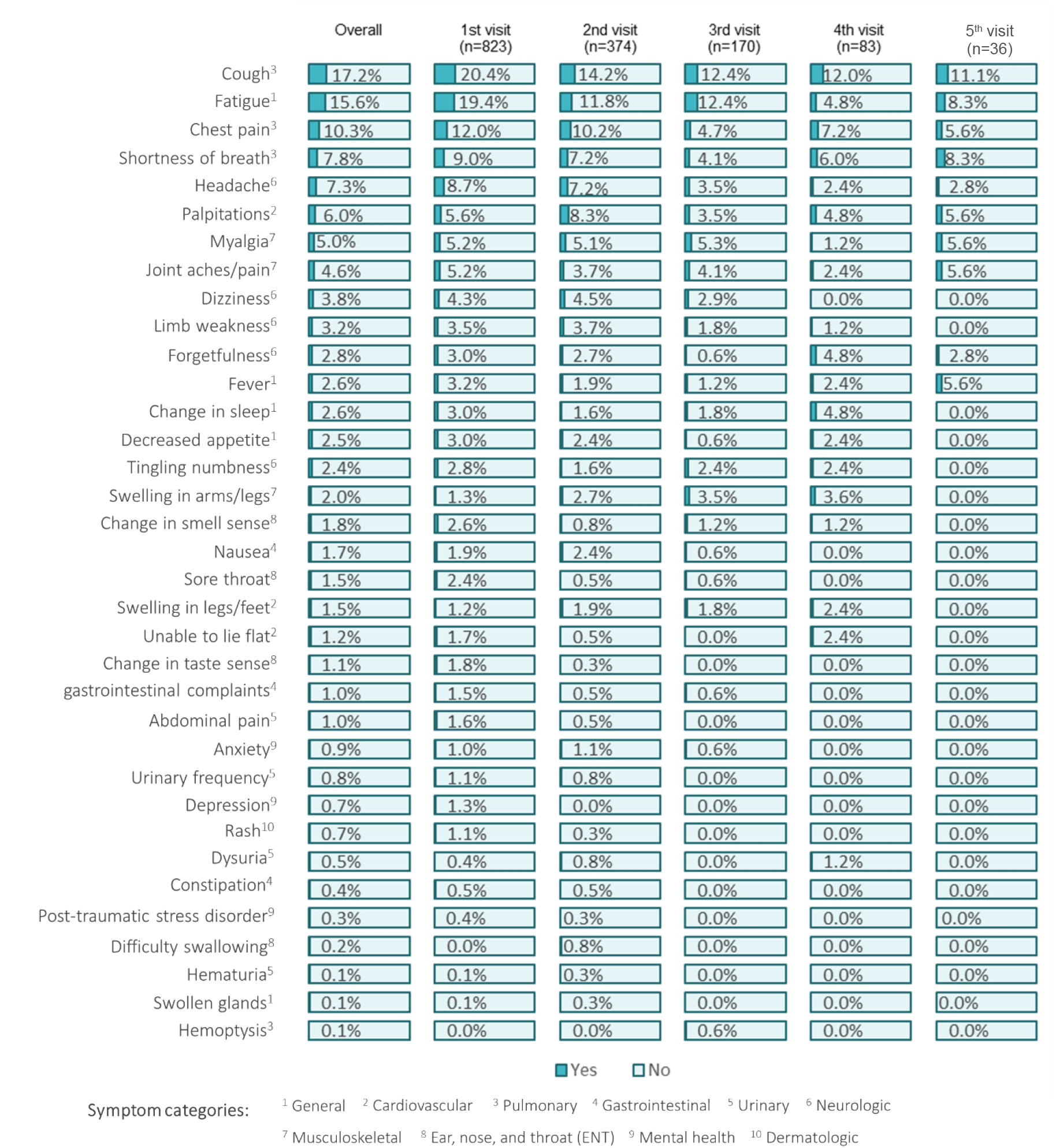
Frequency of long COVID symptoms by PAC-19 clinic visit in Zambia, Aug-20 to Dec-22

In this study, the total person-time at risk was 38,162 person-days. Assuming constant risk, the overall incident rate of COVID-19 symptoms resolving was 12.2 per 1,000 person-days. The resolution incident rate was higher in the severe acute COVID-19 cohort compared to the mild COVID-19, 17.8 vs 11.6 per 1,000 person-days, respectively. In severe acute COVID-19 cohort, the median survival time of persistent symptoms was 54 (IQR: 37-107) days while it was 31 (IQR: 17-89) days in the mild acute COVID-19 cohort (Fig. 2).

**Fig. 2:**
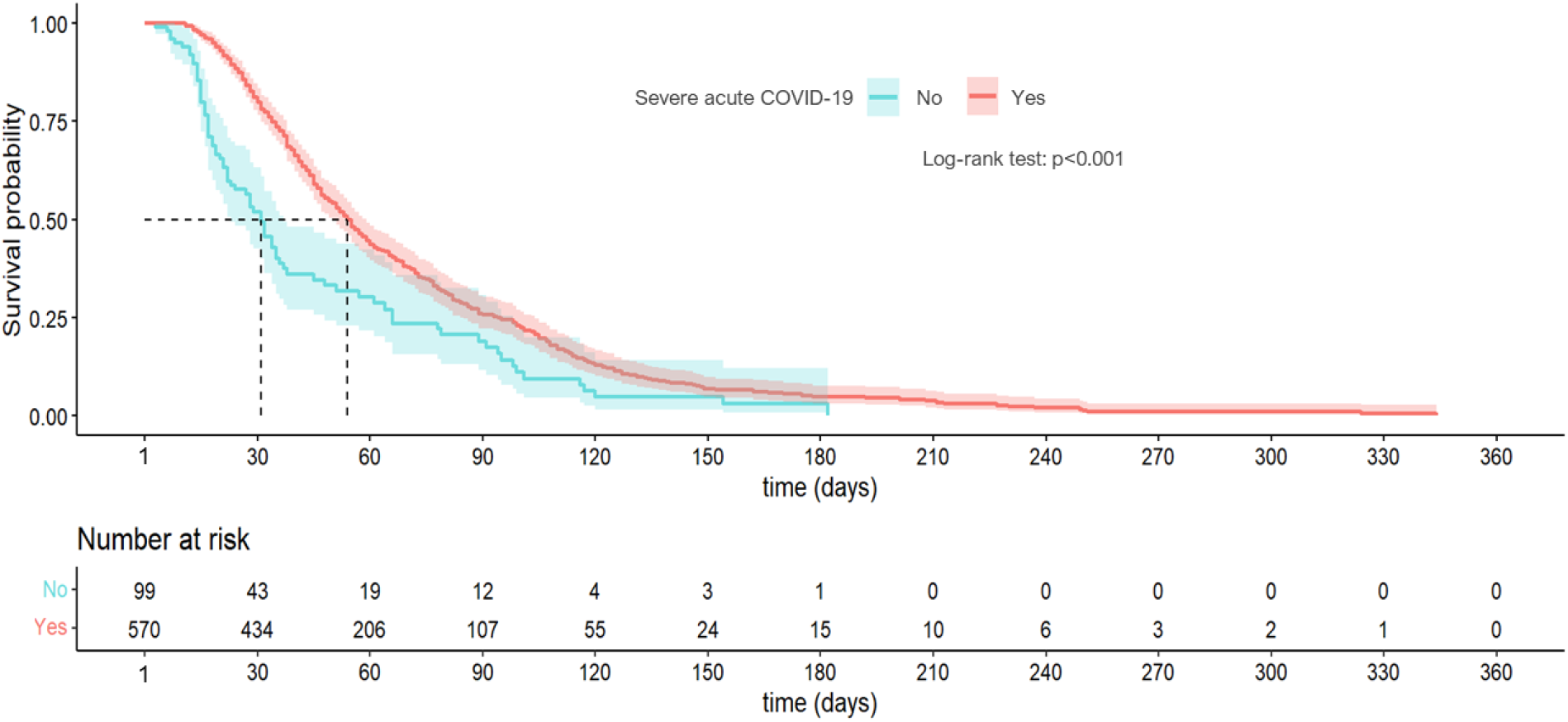
Kaplan-Meier survival curve for post-acute COVID symptoms resolving by severe acute COVID-19 status, Aug-20 to Dec-22

From Cox proportional hazard models, having severe acute COVID-19 disease was associated with reduced hazard of persistent COVID-19 symptoms resolving (adjusted hazard ratio [aHR]: 0.65; 95% confidence interval [CI]: 0.41-0.88). Participants diagnosed with COVID-19 during the Omicron variant dominant phase had 4.05 (95% CI: 1.27-12.9) increased hazard of symptoms resolving compared to those diagnosed during the Alpha wave. Participants vaccinated during the time of PAC-19 clinic follow up had a reduced risk of symptoms resolving (aHR: 0.25; 95% CI: 0.09-0.71) while prior vaccination was associated with increased non-significant hazard of symptom resolving. Females compared males (p=0.391) and age group (p>0.05) were associated with non-significant hazard of persistent COVID-19 symptoms resolving.

The temporal dynamics of persistent COVID-19 symptoms resolving was not constant but may have followed a generalized gamma distribution, i.e., increasing then decreasing (non-monotonic) underlying hazard rate with a peak at 20 days (Fig 3). At peak hazard rate, there was a 2.14% chance that a participant would have their symptoms resolve in the next person-day interval, assuming they had not yet resolved. Cumulatively, 81 (9.8%) participants already had their symptoms resolved at 20 days and symptoms survival probability was at 88.3% (95% CI: 85.9-90.7%). In this parametric AFT model, the effect of the baseline comorbidities was to lengthen (decelerate) the time to symptoms resolving by a time ratio of 86% (adjusted time ratio [aTR]: 1.86; 95% CI: 1.60-2.17) – Table 2. Similarly, hospitalization for ≥15 days during acute COVID-19 significantly lengthened time to symptom resolving by an aTR of 1.52 (95% CI: 1.19-1.93). Prior vaccination accelerated (shortened) the time to symptom resolving by about half (aTR: 0.5, p=0.033) while vaccination during PAC-19 clinic follow up lengthened (decelerated) the time to symptom resolving by 1.97.

**Table 2:** Factors associated with time-to-resolution of persistent COVID-19 symptoms in Zambia, Aug-20 to Dec-22.

| Variables | Stratified Cox Proportional Hazard Model |  |  |  | Parametric AFT Model <sup>1</sup> |  |
| --- | --- | --- | --- | --- | --- | --- |
|  | Unadjusted HR (95% CI) | p-value* | Adjusted HR (95% CI) | p-value* | Adjusted TR (95% CI) | p-value* |
| **Sex** |  |  |  |  |  |  |
| Male | Referent | - | Referent | - | Referent | - |
| Female | 0.93 (0.74-1.17) | 0.543 | 0.90 (0.70-1.15) | 0.391 | 1.08 (0.95-1.23) | 0.257 |
| **Age group (years)** |  |  |  |  |  |  |
| ≤29 | Referent | - | Referent | - | Referent | - |
| 30-39 | 0.58 (0.30-1.12) | 0.103 | 1.13 (0.56-2.29) | 0.737 | 0.93 (0.63-1.37) | 0.723 |
| 40-49 | 0.63 (0.35-1.14) | 0.125 | 1.20 (0.64-2.24) | 0.777 | 0.88 (0.62-1.24) | 0.723 |
| 50-59 | 0.71 (0.40-1.26) | 0.239 | 1.26 (0.68-2.34) | 0.464 | 0.76 (0.54-1.08) | 0.124 |
| 60+ | 0.56 (0.32-0.98) | **0.043** | 1.42 (0.77-2.62) | 0.262 | 0.80 (0.57-1.12) | 0.193 |
|  | Unadjusted HR<br>(95% CI) | p-value* | Adjusted HR<br>(95% CI) | p-value* | Adjusted TR<br>(95% CI) | p-value* |
| **Dominant SARS-CoV-2 variant at diagnosis<sup>2</sup>** |  |  |  |  |  |  |
| Alpha | *Referent* | - | *Referent* | - | *Referent* | - |
| Beta | 1.27 (0.68-2.36) | 0.458 | 1.89 (0.61-5.89) | 0.410 | 0.91 (0.53-1.54) | 0.715 |
| Delta | 1.10 (0.64-1.89) | 0.725 | 2.51 (0.85-7.47) | 0.097 | 0.94 (0.57-1.56) | 0.821 |
| Omicron | 3.58 (1.88-6.80) | **<0.001** | 4.05 (1.27-12.9) | **0.018** | 0.84 (0.48-1.46) | 0.538 |
| **Presence of comorbidities <sup>3,4</sup>** |  |  |  |  |  |  |
| No | *Referent* | - | - | - | *Referent* | - |
| Yes | 0.64 (0.51-0.81) | **<0.001** | - | - | 1.86 (1.60-2.17) | **<0.001** |
| **Hospital length of stay (days) <sup>4</sup>** |  |  |  |  |  |  |
| 1-3 | *Referent* | - | *Referent* | - | *Referent* | - |
| 4-7 | 0.48 (0.31-0.73) | **<0.001** | - | - | 1.01 (0.80-1.26) | 0.952 |
| 8-14 | 0.36 (0.24-0.56) | **<0.001** | - | - | 1.09 (0.86-1.38) | 0.463 |
| ≥15 | 0.15 (0.10-0.23) | **<0.001** | - | - | 1.52 (1.19-1.93) | **0.001** |
| **Severe acute COVID-19<sup>5</sup>** |  |  |  |  |  |  |
| No | *Referent* | - | *Referent* | - | *Referent* | - |
| Yes | 0.40 (0.26-0.61) | **<0.001** | 0.65 (0.41-0.88) | **0.009** | 1.27 (1.12-1.70) | **0.003** |
| **Vaccination status <sup>6</sup>** |  |  |  |  |  |  |
| Unvaccinated | *Referent* | - | *Referent* | - | *Referent* | - |
| Prior vaccination | 1.48 (0.60-0.82) | 0.393 | 1.42 (0.62-3.26) | 0.409 | 0.50 (0.26-0.94) | **0.033** |
| Vaccination during follow up | 0.22 (0.08-0.67) | **0.007** | 0.25 (0.09-0.71) | **0.009** | 1.97 (1.18-3.26) | **0.009** |

**Abbreviation:** AFT – Accelerated failure time; HR - Hazard ratio; and TR – Time ratio for Generalized Gamma distribution; CI – confidence interval.
\* Bolded p-values are significant at p<0.05
<sup>1</sup> Generalized gamma probability distribution parametric model
<sup>2</sup> Dominant SARS CoV-2 variants classification based on GISAID (2023) rather than sequenced specimens from study patients
<sup>3</sup> Pre-existing comorbidity: hypertension, diabetes, cardiovascular disease, cancer, immunosuppression, chronic lung, kidney, and liver diseases, obesity, HIV and TB.
<sup>4</sup> Stratifying variable for the stratified Cox Proportional Hazard model
<sup>5</sup> Composite variable for acute COVID-19 episodic that required supplemental oxygen therapy, intensive care unit stay or treatment with steroids/remdesivir
<sup>6</sup> Vaccination status was a time-varying covariate, i.e., a proportion of patients initially not vaccinated got vaccinated during follow up time.

**Fig. 3:**
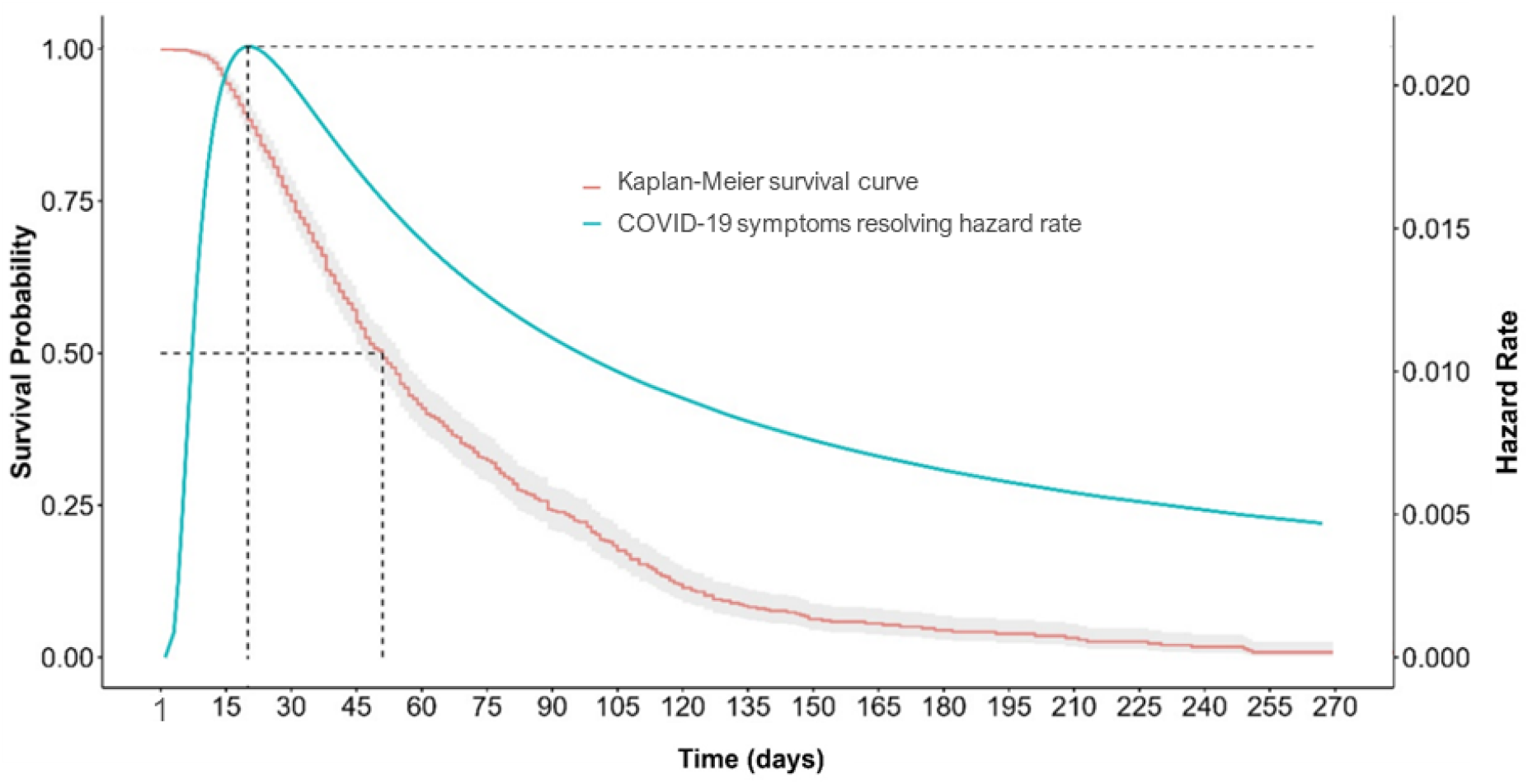
Kaplan-Meier survival curve and generalized gamma probability distribution hazard rate for persistent COVID-19 symptom resolving, Aug-2020 to Dec-2022

## DISCUSSION

Factors associated with the risk of COVID-19 symptoms resolving included severe acute COVID-19, diagnosis with SAR-CoV-2 infection during an Omicron variant dominant period and being vaccinated prior to COVID-19 infection similar to findings in previous studies [36-40]. Exposure to severe acute COVID-19 and vaccination against COVID-19 during follow up were associated with reduced risk of symptoms resolving. Prior vaccination and diagnosed with SARS-CoV-2 during an Omicron variant dominant period had an increased risk of symptoms resolving. Furthermore, the temporal dynamics of post-acute COVID-19 symptoms resolving was not constant but followed an increasing then decreasing trajectory with a peak rate at under one month and a median survival time occurring a month later. The changes in hazard rate over time is consistent with a report on the survival time dynamics of long-lasting symptoms after acute COVID-19 [41].

The median survival time in this study is consistent with findings from another studies in Zambia [15, 27, 29]. This may mean that prolonged morbidity due to COVID-19 potentially impacted on healthcare resource demand post the acute phase of SARS-CoV-2 infections. Simply put, as much as half of the PAC-19 clinic patients were still experiencing lingering symptoms beyond 2 months. Whereas only <1% (1,238) of the over 349,000 confirmed COVID-19 cases presented in PAC-19 clinics, patients with persistent COVID-19 symptoms may have added to the overall burden on healthcare systems. This underlines the utility of specialized post-acute COVID-19 clinics and their necessity to provide continued support during long-term COVID-19 recovery.

The incident rate of symptoms resolution in this cohort was lower than was found in other studies conducted during acute COVID-19 [29, 42, 43]. A lower symptoms resolution incident rate may be related to tissue injury that potentially leads to long-term organ dysfunction and a longer recovery process [3]. Inflammation and immune dysregulation, which is also common in SAR-CoV-2 infections, may have also prolonged recovery and symptoms resolution rate [44]. Furthermore, post-acute COVID-19 symptoms are multi-systemic conditions with long-lasting health effects and thus likely take more time to resolve.

Severe acute COVID-19 was associated with greater persistent symptoms than mild COVID-19 [45]. This is thought not only to be related to the inflammation of the central nervous system but to intensive medical treatments in severe COVID cases which in turn potentially affect the immune response and the body’s ability to recover [46, 47]. Persistent symptoms may furthermore lead to psychological symptoms such as anxiety, depression, and post-traumatic stress disorder which are reported in long COVID patients [48, 49]. These may have adversely contributed to the overall ability of the body to recover.

In this study, vaccination during follow up had reduced hazard of symptoms resolving consistent with another study on prolonged symptoms in people that receive COVID-19 vaccines when already infected with a SARS-Cov-2 virus [50]. This may be related to the overlap of immune processes, potentially leading to immune dysregulation or complications in symptom recovery [51]. In our study, only about one-sixth of the participants were vaccinated prior to SARS-CoV-2 infection and these had an accelerated (shorter) time to persistent symptom resolving. Receiving COVID-19 vaccines prior to SARS-CoV-2 infection is known to enhance vaccine effectiveness by priming the immune system and allowing for a more effective response [52-54]. This thus highlights the importance of timing COVID-19 vaccines primarily way before a SARS-CoV-2 infection to accord the body time to mount an optimal immune response.

The findings that patients diagnosed with COVID-19 during an Omicron variant dominant period had an increased risk of symptom resolution is consistent with reports that Omicron was less severe than previous lineages [40]. This may be due to immunity occasioned from exposure to previous SARS-CoV-2 variants or vaccination which may have moderated severity of post-acute COVID symptoms [55]. Because of an altered spike protein, the Omicron variant replicates more efficiently in the upper respiratory tract rather than in lungs where more severe respiratory illnesses occur [56]. This may have resulted in less severe lung damage with Omicron compared to other variants and a relatively quicker symptoms resolution.

An underlying increasing then decreasing hazard rate may mean that patients had a higher likelihood of recovering from post-acute COVID-19 symptoms relatively early. This is consistent with what is reported about recovery from COVID-19 in previous studies [15, 27, 29]. A decreasing hazard rate following a peak rate at 20 days, however, may suggest prolonged recovery period for the majority of patients. This information likely may help in treatment planning of long COVID and providing patients with realistic expectations about the timing, likelihood, and patterns of recovery from post-acute COVID-19 symptoms that last more than a month.

The findings in this study are subject to some limitations. Firstly, only patients who were hospitalized during acute COVID-19 and attended post-acute COVID-19 follow up care were included in this study. This may represent some potential limitation as the findings may not be generalizable to patients were outpatients during acute COVID-19, sought care elsewhere for post-acute COVID-19 management, or did not seek care at all. Post-COVID-19 services were relatively limited in Zambia during this project, and many more people were infected and hospitalized with COVID-19 than the number who attended post-COVID-19 clinics. Secondly, patients’ classification of the dominant SARS-CoV-2 variant at time of diagnosis was not based on sequenced specimen. Although the dominant SARS-CoV-2 variants may have accounted for a majority of new COVID-19 cases at the time of diagnosis, other variants were still circulating thereby presenting potential for variant misclassification.

## CONCLUSION

Our study found that persistent COVID-19 symptoms in Zambia had a median survival time of about two months and the peak hazard rate at under a month. The median survival time occurring nearly a month after peak hazard rate may suggest that patients that did not initially improve in the first month of their post-acute infection might have a prolonged recovery period. The time to symptoms resolving was, however, associated with severe acute COVID-19, the dominant SARS-CoV-2 variant at diagnosis, prior vaccination status, the presence of comorbidities and hospital length of stay. These results underscore the complexity and dynamics of the time to persistent COVID-19 symptoms resolving and may likely help in treatment planning and providing patients with realistic expectations about their recovery process.

## Disclaimer

The findings and conclusions in this report are those of the authors and do not necessarily represent the official position of the US Centers for Disease Control and Prevention (CDC) or affiliations institutions of authors or the study funders.

## Data Availability

Data are not currently publicly available but may be obtained by a third party. The data are de-identified participant data, available with permission from the Government of the Republic of Zambia – Ministry of Health (MoH). A request to access the data can be made to the Permanent Secretary - Technical Services, Zambia Ministry of Health, Ndeke House Haile Selassie Ave, P.O Box 30205, Lusaka 10101, Zambia, who together with technical leads will review the request and avail the data. Protocols and statistical analysis information is available as per above.

## SUPPLEMENTARY FIGURES

**Supplementary Fig. 1:**
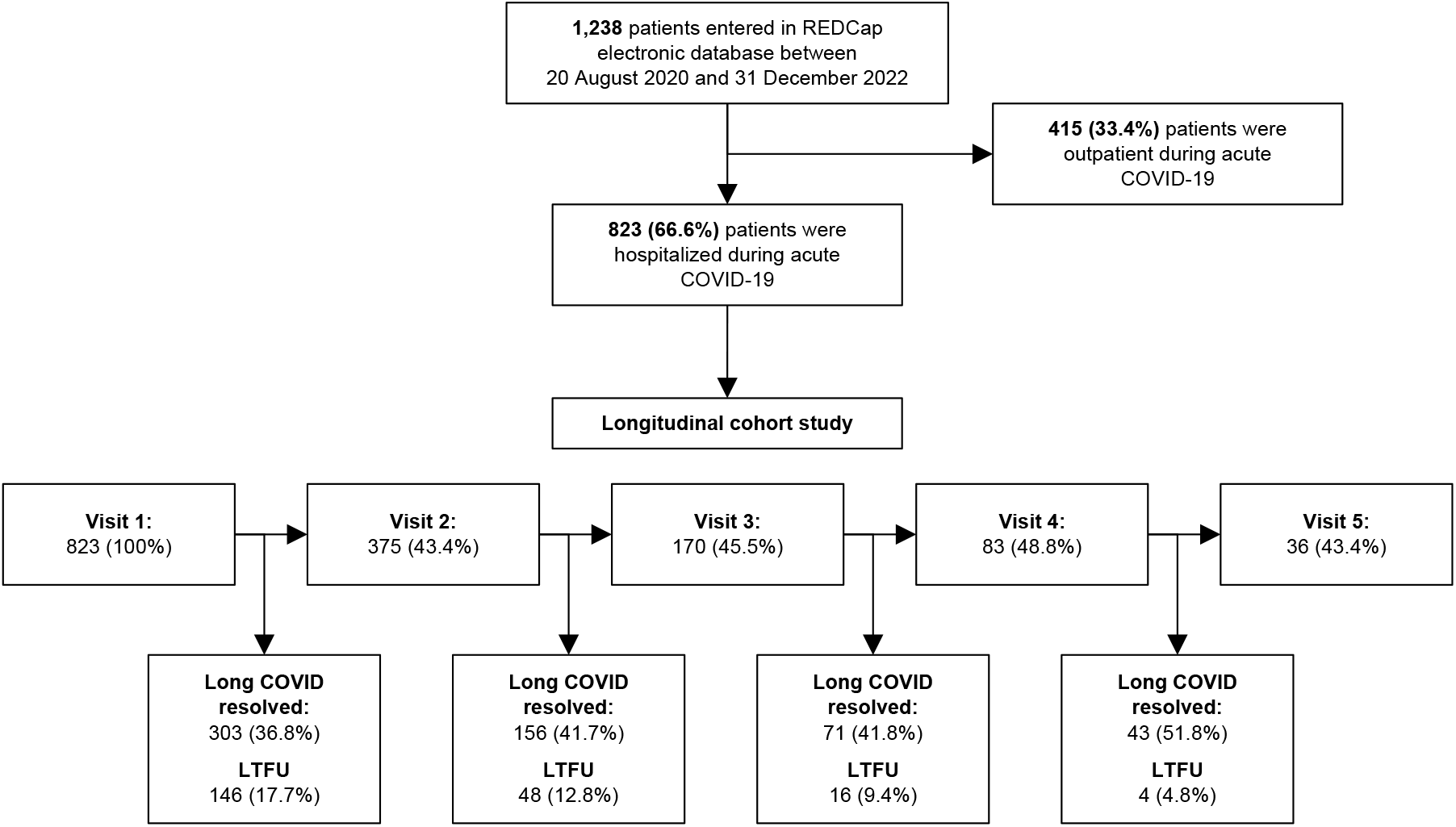
Study analysis flow diagram of participants presenting in PAC-19 clinic in Zambia, Aug. 2020 to Dec. 2022 (LTFU = Lost to follow up)

**Supplementary Fig. 2:**
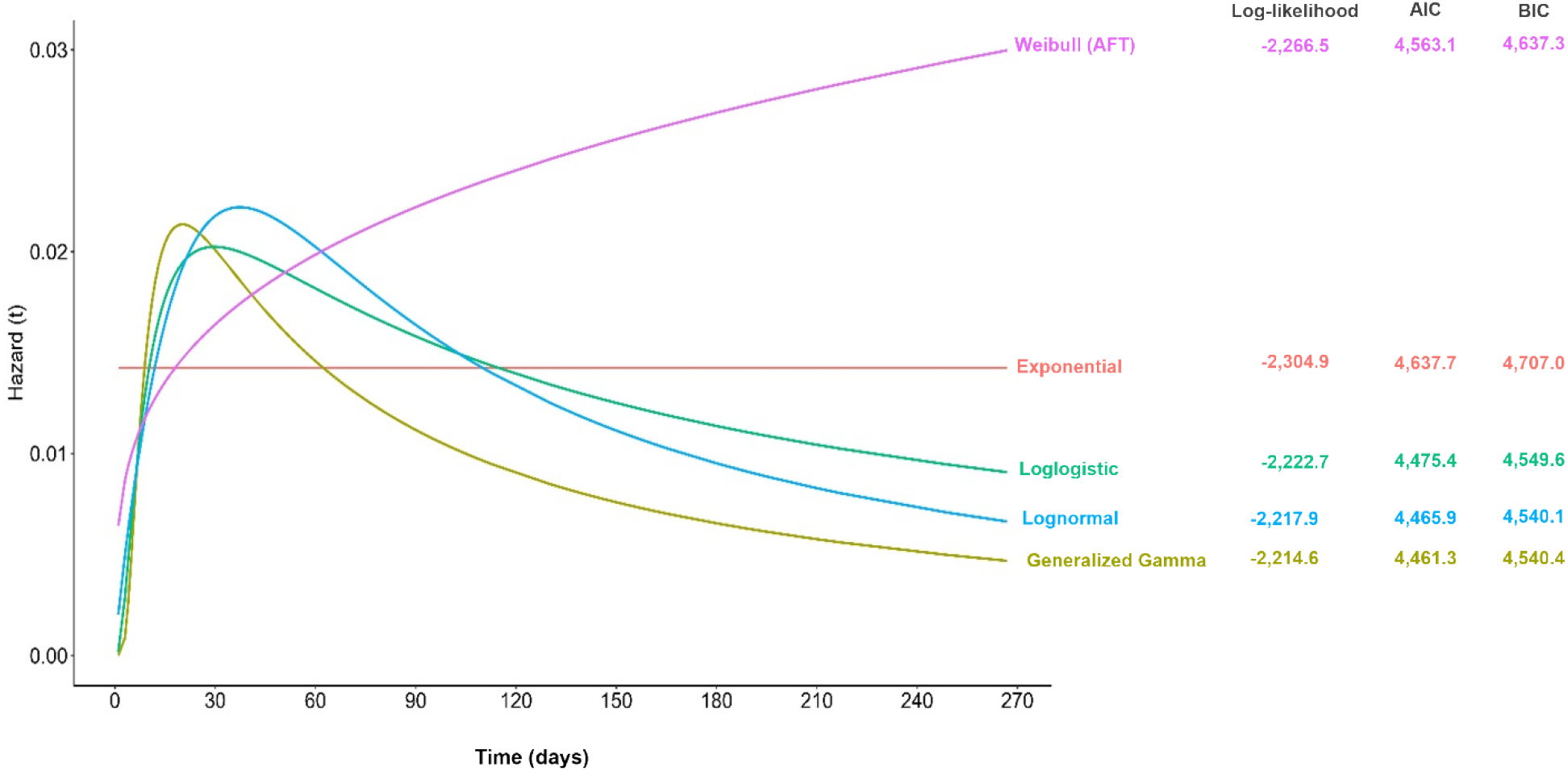
Parametric probability distributions comparison of estimated underlying hazard rate for time to persistent COVID-19 symptoms resolving

